# Effects of low-dose monacolin K on the circulating proteome in individuals with suboptimal cholesterolaemia: A randomised clinical trial

**DOI:** 10.1101/2024.06.19.24309106

**Authors:** Arrigo F.G. Cicero, Patrizia Uboldi, Giangiacomo Beretta, Federica Fogacci, Elisa Grandi, Monika Svecla, Giuseppe Danilo Norata

## Abstract

Red yeast rice (RYR) is a traditional Chinese product obtained by fermenting rice with the yeast *Monascus purpureus* and contains monacolin K, which is chemically identical to lovastatin, a drug with cholesterol-lowering activity. The European Food Safety Authority (EFSA) has evaluated the safety and efficacy of RYR supplements for managing cholesterol levels. In 2018, EFSA published a scientific opinion on the use of monacolin K from RYR in food supplements, concluding that monacolins from RYR may raise significant safety concerns at a use level of 10 mg/ day. Following that, the European Commission declared in 2022 that RYR products must contain less than 3 mg of monacolins for daily consumption. The aim of this work was to perform a comprehensive profiling of plasma markers of muscle and liver dysfunction by extensive untargeted plasma proteomics in healthy volunteers with suboptimal cholesterolaemia who were randomly assigned to receive a dietary supplement containing RYR (total monacolin <3 mg) or placebo for one month.

No changes in classical markers of liver (AST, ALT) or muscle (CPK) function were detected in the plasma samples of patients treated with the supplement compared to placebo. Interestingly, the analysis of circulating proteins marking an early acute response in the liver, such as serum amyloid A4, orosomucoid 2, haptoglobin-related protein, prothrombin, α-1-antitrypsin, α-2-HS-glycoprotein, serum amyloid P (APCS), orosomucoid 1, c-reactive protein (CRP) and α-2- macroglobulin confirmed an overlapping profile in the two groups. Similarly, the analysis of ryanodine receptor 1, titin, dystrophin and myosin 7 again showed a similar profile in the two groups. These data indicate that a low dose of monacolin K (<3 mg/day) in subjects with suboptimal cholesterolaemia does not increase levels of markers of liver and skeletal muscle function in plasma, excluding a deleterious effect of monacolin K on these tissues.

## 1. Introduction

Red yeast rice (RYR) is a traditional Chinese product obtained by the fermentation of rice with the yeast *Monascus purpureus* [1]. RYR contains monacolins, and is particularly enriched with monacolin K, which is chemically identical to lovastatin, a prescription drug with cholesterol-lowering activity [1]. Thus, RYR may represent a natural remedy for people with moderately elevated cholesterol levels. A meta-analysis of randomised controlled trials has shown that RYR supplementation significantly reduces total cholesterol, LDL-C and triglycerides [2]. In terms of safety, available information on potential adverse effects indicates that individuals taking monacolin k at a dose of 10 mg/day may experience muscle symptoms or liver dysfunction and or severe acute hepatitis; serious adverse events have been reported even at a dose of 3 mg/day [3].

The European Food Safety Authority (EFSA) has evaluated the safety and efficacy of RYR supplements for managing cholesterol levels. In 2018, EFSA published a scientific opinion on the use of monacolin K from RYR in food supplements and concluded that monacolins from RYR may raise significant safety concerns at a use level of 10 mg/ day. [4]. For this reason, the European Commission declared in 2022 that RYR products must contain less than 3 mg of monacolins for daily consumption [5].

Interestingly, an extensive analysis of adverse event reporting systems (FAERS and CAERS) and available case reports has shown that the occurrence of rhabdomyolysis or severe acute hepatitis that could be associated with the use of RYR appears to be extremely rare compared to the occurrence with statins (which is rare to common) [6].

In terms of all musculoskeletal disorders, 363,879 cases were reported in the FAERS from September 2013 (when the first case related to RYR use was recorded) to 30 September 2023, with cases associated with RYR use being very low and accounting for 0.008% of cases. During the same period, 27,032 cases of hepatobiliary disorders were reported, with cases attributable to RYR use accounting for 0.01% of all cases. A low rate of muscle symptoms and liver dysfunction attributable to RYR use was also observed in the CAERS database. This profile mirrors that of meta-analyses of randomised clinical trials of RYR, in which RYR use was not associated with either liver dysfunction or muscular adverse symptoms [7].

A current gap in the literature is the evaluation of the safety of RYR extracts containing monacolin K <3 mg. To this end, we performed comprehensive profiling of plasma markers of musculoskeletal and liver dysfunction by extensive, untargeted plasma proteomics in healthy volunteers with suboptimal cholesterolaemia randomised to receive a dietary supplement (NUT) containing RYR (total monacolin <3 mg) or placebo.

## 2. Material and Methods

### 2.1 Study design

Individuals were enrolled in the Outpatient Service of the Cardiovascular Medicine Unit by the IRCCS University Hospital of Bologna of the Medical and Surgical Sciences Department (DIMEC) of the University of Bologna. After screening 46 healthy adults with suboptimal cholesterolaemia, 40 volunteers were enrolled in a double-blind, placebo-controlled, cross-over randomised clinical trial.

Participants with LDL-C levels between 115 and 190 mg/dL were randomised to receive either a dietary supplement (referred to as NUT) containing RYR (total monacolin <3 mg) or a placebo, both in combination with a standard Mediterranean diet (rich in vegetables, whole grain carbohydrates, extra-virgin olive oil, and low in salts, processed foods and animal fats) with low cholesterol content (Standard of Care - SOC), (<200 mg/day, as indicated by the European Atherosclerosis Society guidelines).

All patients received an information document describing the study and signed a consent form to participate in the study. The study was approved by the Ethics Committee of the University of Bologna and registered on www.clinicatrial.gov (ID: NCT06368258).

#### Inclusion criteria

- Male or female aged ≥ 30 years and ≤ 70 years.
- Body mass index between 18 Kg/m^2^ and 35 Kg/m^2^
- Total cholesterol level between 200 and 280 mg/dL and/or LDL-C between 130 mg/dL and 190 mg/dL.
- TG<400 mg/dL.
- Subjects who, have a low or moderate cardiovascular risk according to the SCORE charts (defined as total cardiovascular risk < 5%) [8] and for whom the intervention strategy does not require a pharmacological lipid-lowering intervention, according to the 2019 ESC/EAS guidelines.
- Subjects who are able to communicate, make themselves understood and comply with the requirements of the study.
- Subjects who agree to participate in the study and have dated and signed the informed consent form.

#### Exclusion criteria

- Ongoing lipid lowering therapy;
- Ongoing drug treatment that has not been stabilised for at least 3 months;
- Known current thyroid, gastrointestinal or hepatobiliary disease (including liver transaminases ≥3ULN) and any muscular disorders (even subclinical, including serum CPK ≥3ULN);
- Current or previous alcohol abuse;
- Pregnancy and breastfeeding
- Known previous intolerance to RYR
- History or clinical evidence of significant concomitant disease that could compromise the safety of the subject or ability to complete the study;
- Any medical or surgical condition that would limit the patient’s compliance with the study protocol.

The timeline of the study is summarised in Figure 1.

**Figure 1.**
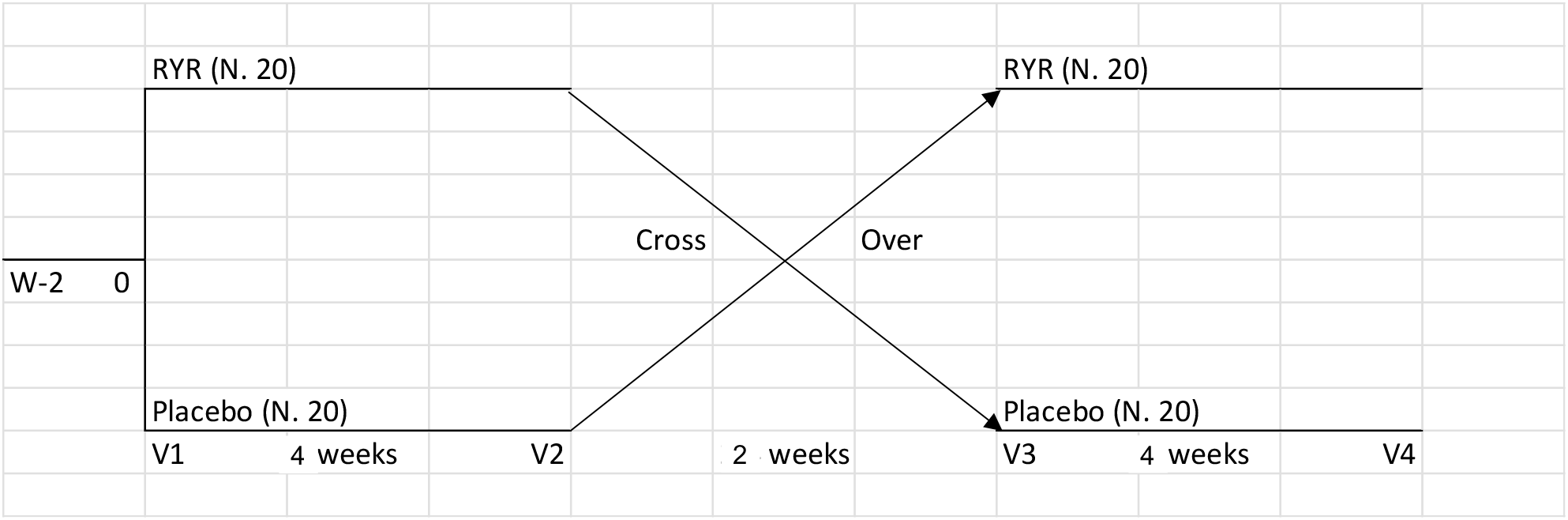
**Study design.**

All subjects who met the inclusion criteria were prescribed the SOC diet for a period of 14 days (lipid stabilisation period - LSP) upon inclusion in the study.

At the end of this run-in period, subjects whose LDL-C levels were reduced by less than 10% and with a compliance to the diet higher than 80% were randomised to receive either NUT or PLACEBO (1 tablet of either preparation once-daily at evening meal) for 6 weeks (± 5 days). After a 2 week washout period, participants were assigned to the alternative treatment (crossover).

The tested product (NUT) and the placebo were provided free-of-charge from MEDA S.p.A. (Monza, Italy). Study-specific insurance was requested and paid from the University funds of the principal investigator.

During the study, subjects were examined during 5 visits:

#### • **Screening visit** (-2 weeks ± 5 days)

After obtaining informed consent, the investigator performed a physical examination to record vital signs (weight, blood pressure, cardiac rhythm, waist circumference) and medications and evaluated the presence of metabolic syndrome criteria.

A urine test was performed in women of fertile age to exclude pregnancy. None experienced pregnancy during the trial.

#### · Enrolment, randomisation and study visits (Time 0)

A study-specific blood sample was taken (under fasting conditions) and the following analyses were performed: LDL-C, total cholesterol, HDL-C, triglycerides, plasma glucose, ALT, AST, gamma-GT, CPK followed by an extensive proteomic analysis.

All participants were randomised according to the randomisation list of the respective centre, and received a pack (5 blisters, 50 tablets in total) of either NUT or PLACEBO. Participants were examined by a physician to record their vital signs (weight, height, blood pressure, cardiac rhythm, waist circumference).

The same analyses were repeated at weeks 6, 8 and 12

### 2.2. Assessments

#### 2.2.1. Clinical data and anthropometric measurements

Subjects’ personal history was evaluated with special attention to cardiovascular and metabolic diseases, assessment of dietary and smoking habits (both with validated semi-quantitative questionnaires) [9], physical activity, and pharmacological treatments.

Waist circumference (WC) was measured at the end of a normal expiration, in a horizontal plane at the midpoint between the inferior margin of the last rib and the superior iliac crest. Height and weight were measured to the nearest 0.1 cm and 0.1 kg, respectively, with subjects standing erect with eyes directed straight wearing light clothes and with bare feet. BMI was calculated as body weight in kilograms, divided by height squared in meters (kg/m^2^).

#### 2.2.2. Blood pressure measurements

Systolic and diastolic blood pressure measurements were performed in each subject, supine and at rest, using a validated oscillometric device, with a cuff of the appropriate size applied on the right upper arm. To improve detection accuracy, three blood pressure (BP) readings were sequentially obtained at 1-min intervals. The first one was discarded, and the average between the second and the third was recorded [10].

#### 2.2.3. Laboratory Data

Biochemical analyses were performed on venous blood, withdrawn after overnight fasting (at least 12 hours). Plasma was obtained by adding disodium ethylenediaminetetraacetate (Na2EDTA) (1 mg/mL) and blood centrifugation at 3000 RPM for 15 min at room temperature.

Trained personnel performed laboratory analyses immediately after centrifugation according to standardised methods [11], to assess TC, HDL-C, TG, Apo-B, apolipoprotein A1 (Apo-A1), fasting plasma glucose (FPG), creatinine, estimated glomerular filtration rate (eGFR) and liver transaminases. LDL-C was obtained by the Friedewald formula. Non-HDL-C was calculated from the difference between TC and HDL-C.

#### 2.2.4. Safety and Tolerability

Safety and tolerability were evaluated through continuous monitoring of any adverse event, clinical safety, laboratory findings, vital sign measurements, and physical examinations.

#### 2.2.5 Blood collection

Blood samples (3 ml) were collected in EDTA tubes. The plasma was isolated following ultracentrifugation and an aliquot of 500 µl was stored at -80°C. After obtaining all samples, 20 µl of plasma was mixed with 40 µl of ammonium bicarbonate 50mM (final pH=8.5). Proteins were reduced by incubation with 3 µl DTT 100 mM, for 30 minutes at 55°C. Protein alkylation was then performed at room temperature, by incubating with 6 µl of iodoacetamide 150mM for 20 minutes in the dark. Trypsin digestion (enzyme to protein ratio 1:20) was performed overnight at 37°C and stopped by acidification with trifluoroacetic acid (final percentage 1%).

### 2.3 Liquid chromatography-mass spectrometry (LC-MS/MS) analysis of plasma proteome

An Ultimate 3000 nano-LC system (Thermo Fisher Scientific, Watlham, MA) connected to an Orbitrap Fusion^TM^ Tribrid™ Mass spectrometer (Thermo Fisher Scientific) equipped with a nanoelectrospray ion source was used. Peptide mixtures were pre-concentrated into an Acclaim PepMap 100 – 100µm x 2cm C18 (Thermo Fisher Scientific) and separated on EASY-Spray column ES802A, 25 cm x 75 µm ID packed with Thermo Scientific Acclaim PepMap RSLC C18, 3 µm, 100 Å using mobile phase A (0.1 % formic acid in water) and mobile phase B (0.1% aqueous formic acid /acetonitrile (2:8)) with the following elution gradient: 4-28% for 90min, 28-40% for 1min, followed by 95% for a total runtime of 150min, at a flow rate of 300 nL/min. The temperature of the column was set to 35°C and the sample was injected four times. The injection volume was 3 µL for each sample. Two blanks were run between samples to prevent sample carryover. MS spectra were collected in positive ion mode over a m/z range of 375 – 1500 Da (resolution 120,000), automatic gain control (AGC) target 4 × 105, maximum injection time of 50 ms, operating in the data-dependent mode, cycle time 3 sec between master scans. MS/MS spectra were collected in centroid mode. HCD was performed with collision energy set at 35 eV.

### 2.4 Proteomic Data Analysis

Protein abundance was calculated with the generation of spectral features by the node FeatureFinderMultiplex followed by PIA-assisted FDR-multiple scores estimation and filtering (combined FDR score<0.01), their ID mapping and combination with peptide IDs, their subsequent alignment, grouping and normalisation (e.g., MapAlignerIdentification, FeatureUnlabeledQT and ConsensusmapNormalizer nodes). DAVID (The Database for Annotation, Visualization, and Integrated Discovery) platform was used for gene ontology (GO) enrichment analysis. GO was performed in our protein LFQ dataset and significant terms associated with the gene set were selected based on the FDR<0.05. Gene set enrichment analysis (GSEA) was performed with ClusterProfiler33 on a pre-ranked gene list for Gene Ontology terms. Gene sets with size <3 and >500 genes were excluded and enriched gene sets with FDR<0.05 were considered significant. Graph Pad- Prism8 and Excel were used for graphic presentation and statistical analysis. For the integration of the proteome for functional analysis, QIAGEN’s Ingenuity® Pathway Analysis (IPA®, QIAGEN) was used.

### 2.5 Statistical analysis

Based on the literature, 15 patients per intervention group (total 30 patients) were required to have a 90% probability of observing a difference of 8% in the reduction of LDL-C from baseline to week 6 between the two treatment groups with level of significance of 5% (two-tails test). Taking into account a possible drop-out rate and/or non-compliance to the protocol of 25%, 20 patients were randomized in each intervention arm (40 subjects in total).

Data were analysed using intention to treat by means of the Statistical Package for Social Sciences (SPSS) version 27.0 (IBM Corporation, Armonk, NY, USA) for Windows.

A complete descriptive analysis of the collected parameters was carried out as per protocol. Categorical variables were expressed as absolute number and percentage and compared with the Fisher corrected chi-square test or the Wilcoxon-Rank test, based on whether they were nominal or ordinal. Continuous variables were expressed as mean ± standard deviation (SD) or mean and standard error (SEM), and compared by analysis of Covariance (ANCOVA) followed by post-hoc Tukey test or by Kruskal-Wallis non parametric analysis of variance followed by Dunn’s pairwise test, depending on whether their statistical distribution was normal or not. To verify the basic assumption of the cross-over design, the presence of a carryover effect was excluded.

The minimum level of statistical significance was set to *p*< 0.05 two-tailed. The Dixon’s Q test was always performed to exclude extreme values.

Efficacy analyses have been performed considering the intention-to-treat (ITT) population, i.e. all subjects with at least one post-baseline control. A sensitivity analysis of the primary variable was also planned in the per-protocol population, i.e. all subjects without major protocol violations.

## Results

In this report, we present the results of the first part of the clinical trial, in particular the profile of biochemical markers and the proteomic signature between T0 and T1 of subjects in the NUT group compared to placebo, with a focus on safety markers, including liver dysfunction and skeletal muscle alterations. The baseline characteristics of the subjects who participated in the study are shown in Table 1. At T1, no significant differences were observed between the NUT group and the placebo group for AST, ALT, CPK and the other biochemical parameters analysed (Table 2).

**Table 1.** Baseline characteristics.

**Figure 1 – Study design**
|  | NUT | PLACEBO | p |
| --- | --- | --- | --- |
| Age (years) | 57,5 (12,7 ) | 63,1 (16,2) | ns |
| TC (mg/dL) | 217,47 (20,8) | 215,24 (27,7) | ns |
| TG (mg/dL) | 121,9 (50,2) | 111,29 (27,7) | ns |
| HDL-C (mg/dL) | 49,4 (10,1) | 50,2 (13,6) | ns |
| LDL-C (mg/dL) | 143,6 (19,4) | 142,7 (22.0) | ns |
| ApoB (mg/dL) | 102,2 (21,5) | 101,6 (31,1) | ns |
| ApoAI (mg/dL) | 146,7 (26,9) | 150,2 (31,2) | ns |
| FPG (mg/dL) | 90,1 (7,4) | 92,6 (12.0) | ns |
| AST (U/L) | 22,5 (5,6) | 22,9 (4,2) | ns |
| ALT (U/L) | 20,2 (9,6) | 21,8 (8,4) | ns |
| CPK (U/l) | 137,9 (69,3) | 106,2 (58,6) | ns |

**Table 2.** Biochemical parameters at T1.

|  | NUT | PLACEBO | p |
| --- | --- | --- | --- |
| TC (mg/dL) | 216,9 (21,1) | 216,4 (25,4) | ns |
| TG (mg/dL) | 125,3 (66,7) | 131,4 (74,3) | ns |
| HDL-C (mg/dL) | 49,0 (10,3) | 50,2 (9,9) | ns |
| LDL-C (mg/dL) | 142,8 (16,5) | 139,9 (23,7) | ns |
| ApoB (mg/dL) | 99,2 (15,3) | 96,6 (15,4) | ns |
| ApoAI (mg/dL) | 148,5 (25,6) | 147,3 (22,4) | ns |
| FPG (mg/dL) | 87,8 (5,9) | 92,2 (11,2) | ns |
| AST (U/L) | 22,7 (7,9) | 21,6 (5,1) | ns |
| ALT (U/L) | 23,2 (9,6) | 24,4 (11,6) | ns |
| CPK (U/l) | 123,7 (57,6) | 95,6 (64,4) | ns |

To profile the changes in the proteome signature of the different groups, we performed shotgun proteomics on plasma samples collected from each subject at T0 and T1 (Figure 2A). Principal component analysis revealed no specific clustering of samples collected under the different conditions (T0 vs T1, NUT vs placebo), thus excluding drastic alterations in the proteome following the treatment with a supplement containing monacolin K (Figure. 2B). The absence of major changes is also confirmed by the unsupervised hierarchical analysis and k-means clustering which did not distinguish between T0 and T1 of subjects in the NUT groups compared to placebo (Figure 2C).

**Figure 2.**
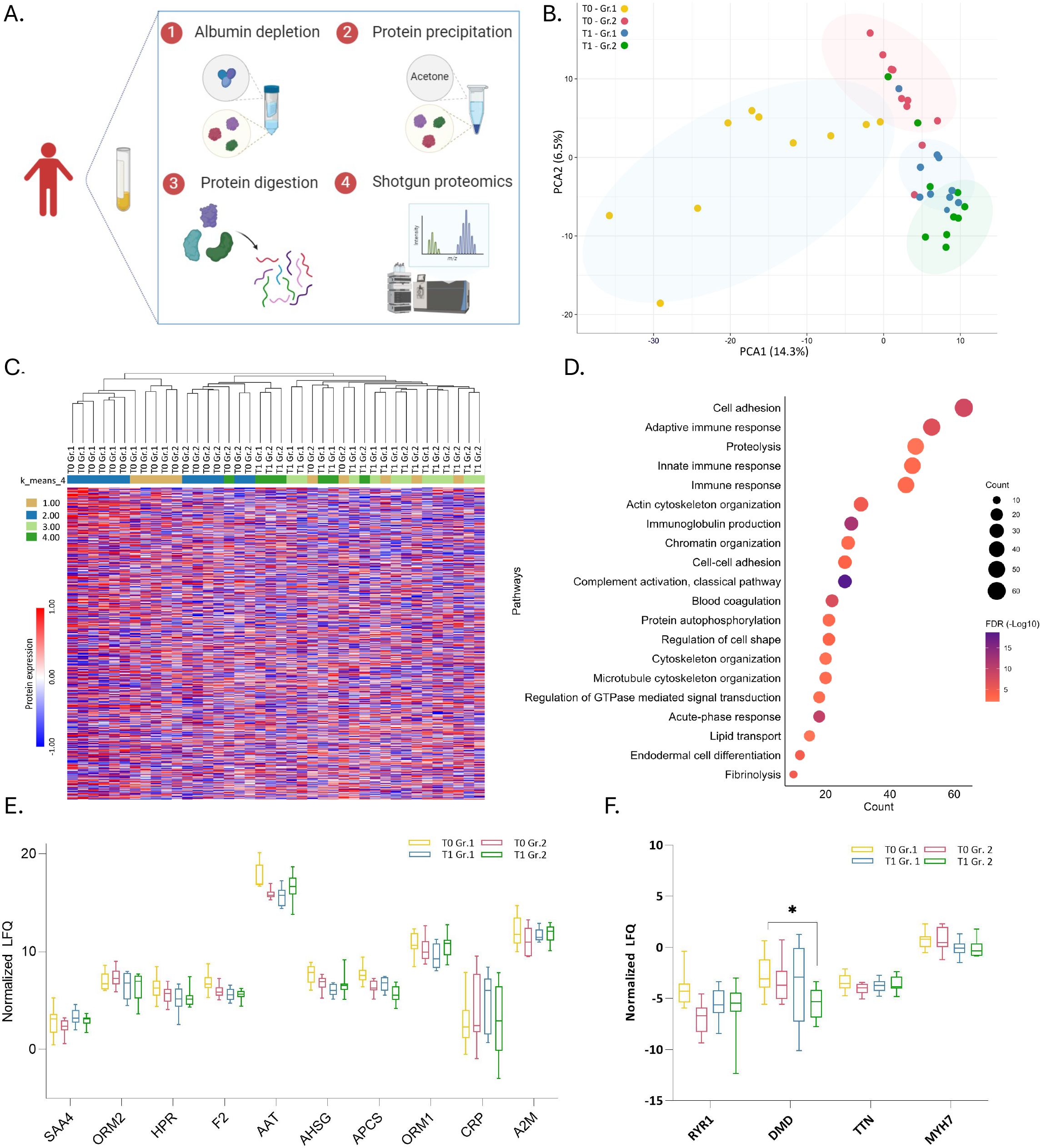
Plasma proteome signature in patients treated with red yeast rice. *(**A**)* Scheme depicting proteomic strategy used to identify and label-free quantify (LFQ) differentially expressed proteins in patients *(**B**)* Unsupervised principal component analysis (PCA) clustering for the total proteome showing the four groups enrolled in the study *(**C**)* Total plasma proteome showing four groups overlapping based on the k-means and hierarchical clustering *(**D**)* Top 20 gene ontology (GO) pathways on the plasma proteome (***E***) Abundance (normalized LFQ) of liver produced proteins associated to acute phase response (***F***) Abundance (normalized LFQ) of plasma proteins related with skeletal muscle. (Gr.1 is placebo and Gr.2 is NUT).

The overall plasma proteome was similar across all subjects at both T0 and T1, with no differences observed between the NUT group (Gr.2) and the placebo group (Gr.1). Most proteins were identified in all subjects at T0 and T1 and did not differ in the NUT group (Gr.2) compared to placebo. Less than <1% of proteins showed significant differences between the NUT and placebo groups. As expected, gene ontology analysis (FDR<0.05) of the total proteome quantified in plasma highlighted several proteins belonging to pathways related to the immune response, acute phase proteins, immunoglobulin production, platelet function, coagulation, complement activation, plasma lipid metabolism (Figure 2D).

Among the proteins identified, those marking liver function such as serum amyloid A4 (SAA4), orosomucoid 2 (ORM2), haptoglobin-related protein (HPR), prothrombin (F2), α-1-antitrypsin (AAT), α-2-HS-glycoprotein (AHSG), serum amyloid P (APCS), orosomucoid 1 (ORM1), c-reactive protein (CRP), α-2-macroglobulin (A2M) presented an overlapping profile in subjects in the NUT group compared to placebo (Figure. 2E). The same analysis was performed for proteins marking skeletal muscle function (Figure 2F) such as Ryanodine receptor 1 (RYR1), Titin (TTN), Dystrophin (DMD), Myosin 7 (MYH7) in plasma which had an overlapping profile in subjects in the NUT group compared to placebo. These data suggest that NUT treatment does not impact the proteomic profile nor of proteins related to liver function neither of those related to skeletal muscle function. No specific expression patterns/differences among samples were detectable when gene ontology analysis for acute phase proteins and actin cytoskeleton organization (Figure 3) or complement activation and adaptive immunity was investigated (Figure 4), again confiming no major differences in the protein expression profile between the different groups.

**Figure 3.**
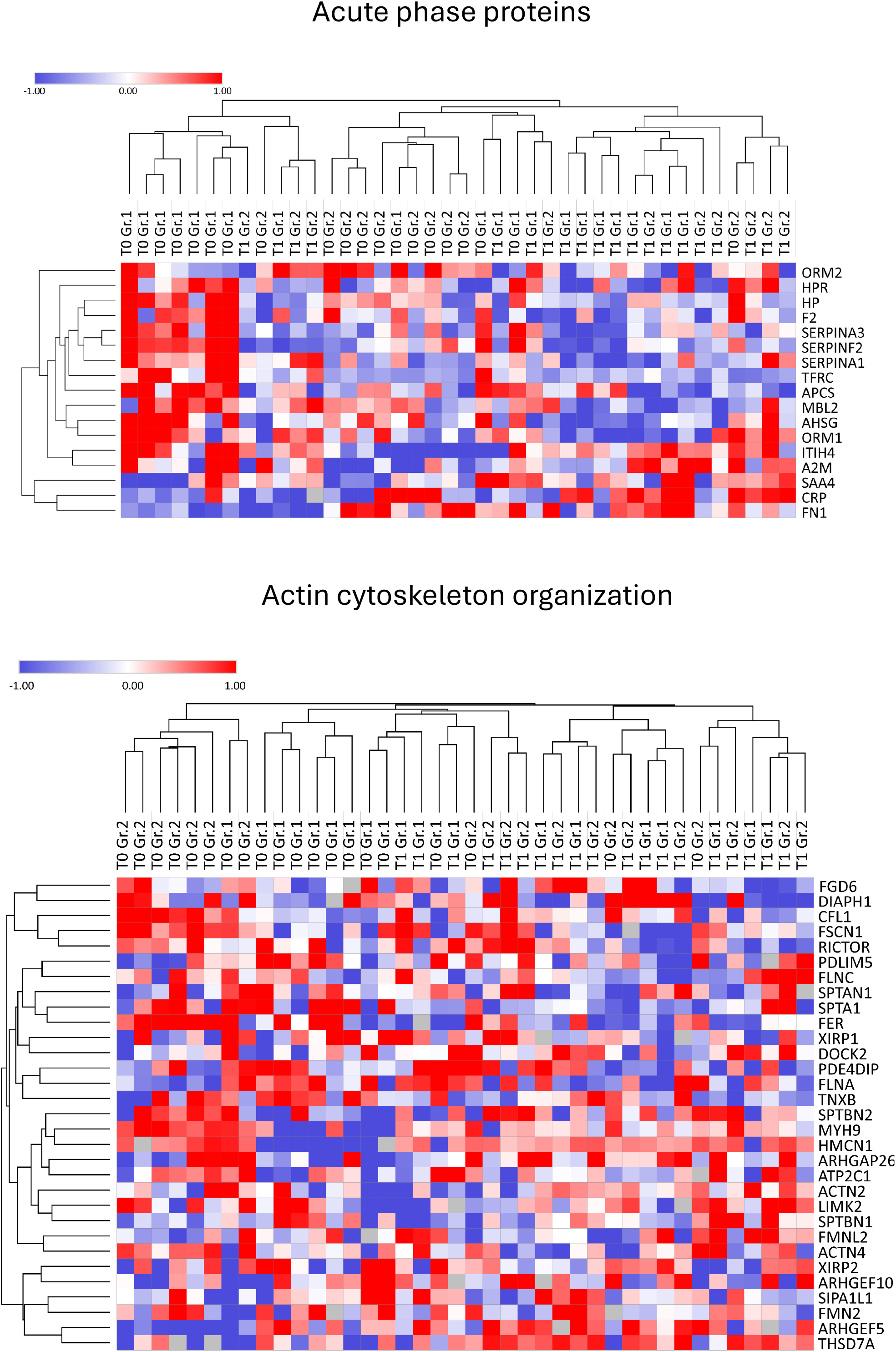
Gene ontology analysis for proteins of the acute phase and of actin cytoskeleton organization. (Gr.1 is placebo and Gr.2 is NUT).

**Figure 4.**
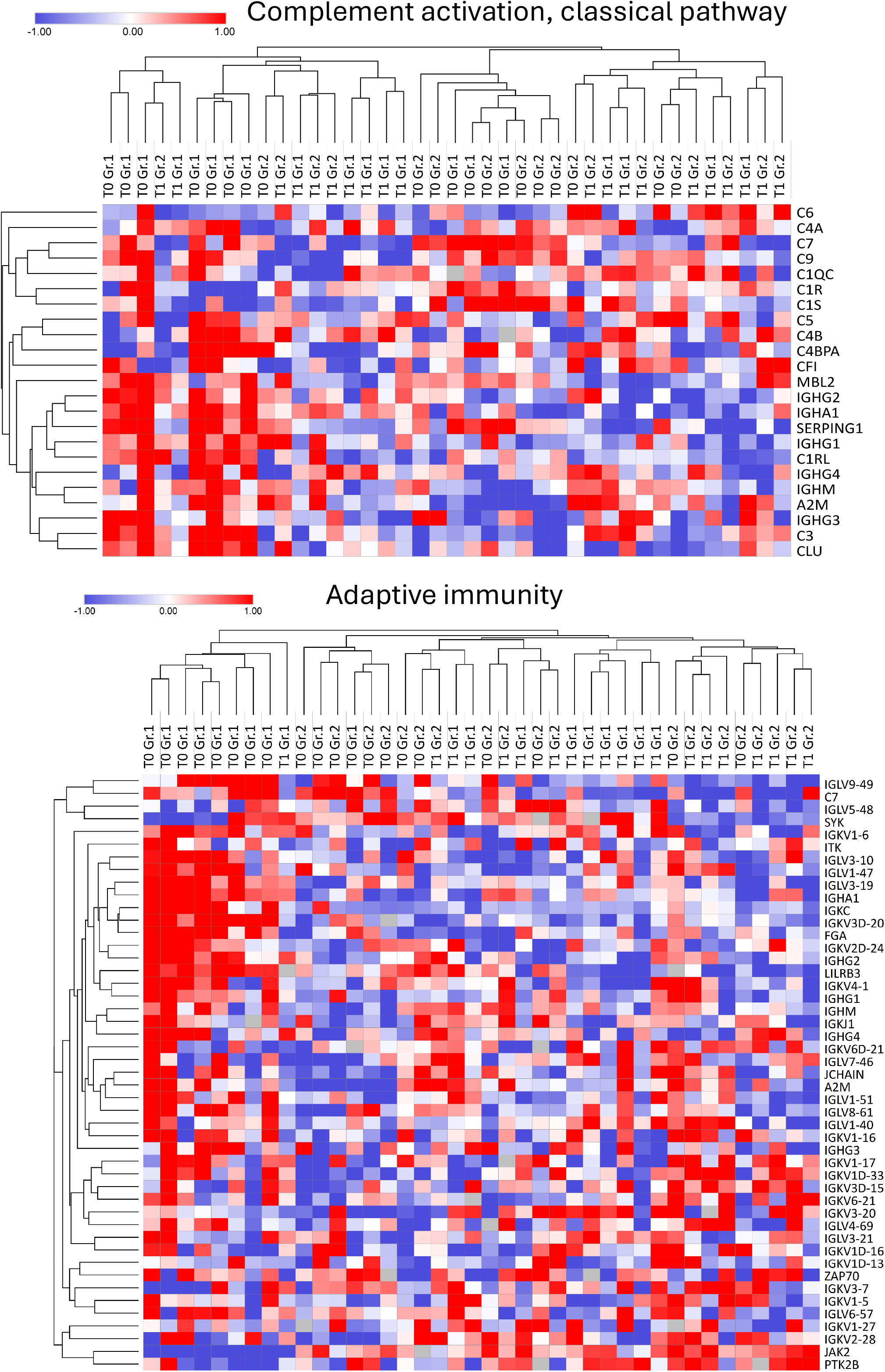
Gene ontology analysis for proteins of the complement pathway and adaptive immunity (Gr.1 is placebo and Gr.2 is NUT).

## Discussion

The aim of this clinical study was to investigate the potential effect of low dose monacolin K (<3 mg/daily) as RYR supplement on the circulating proteome in 40 individuals with suboptimal cholesterolaemia. The study had a crossover design, and each subject received a dietary supplement (referred to as NUT) containing RYR (total monacolin <3 mg) or a placebo, both in combination with a standard Mediterranean diet. One arm was first randomised to placebo and later to NUT, while the second arm was first randomised to NUT and later to placebo. Here we report the results from the first part of the clinical trial in which 20 patients were randomised to receive NUT and 20 patients were randomised to receive placebo. Baseline characteristics were similar between the two groups. The analysis of biochemical parameters 4 weeks after the subjects were randomised to the two arms revealed no differences in key parameters associated with liver function such as AST or ALT and muscle stress such as CPK. Although this analysis already excluded a major impact of RYR containing monacolin K <3 mg/day, we performed an in-depth analysis to monitor changes in early markers of liver and muscle dysfunction by profiling the signature of the plasmatic proteome with untargeted proteomics. The principal component analysis of samples tested at T0 and T1 from the NUT and placebo groups already provided an overview of a very similar plasmatic proteomic profile at baseline and after the treatment, suggesting that the vast majority of proteins that can be detected in plasma were unaffected by the treatment with monacolin K. When we focused our analysis on circulating proteins marking an early acute response in the liver, such as serum amyloid A4, orosomucoid 2, haptoglobin-related protein, prothrombin, α-1-antitrypsin, α-2-HS-glycoprotein, serum amyloid P (APCS), orosomucoid 1, C-reactive protein (CRP) and α-2- macroglobulin their expression profile was almost overlapping between the NUT group and the placebo group as well as between T1 and T0, thus excluding any impact of the treatment with monacolin K (<3 mg/daily) on liver function. Similarly, we focused our analysis on circulating proteins marking skeletal muscle function such as ryanodine receptor 1, titin, dystrophin and myosin 7 and again the profile was almost superimposable between the different groups, further confirming that monacolin K at the dose useddoes not affect muscle function.

In summary, the data obtained from this clinical trial reassure on the safety of low-dose monacolin K (<3 mg/day) in subjects with suboptimal cholesterolaemia, as no changes in markers of liver and skeletal muscle function were observed compared to either baseline or the placebo group.

## Data Availability

All data produced in the present study are available upon reasonable request to the authors

## Acknowledgements

GDN is supported by PNRR Missione 4, [Progetto CN3-National Center for Gene Therapy and Drugs based on RNA Technology], PNRR Missione 4 [Progetto MUSA-Multilayered Urban Sustainability Action], PNRR Missione 6 [PNRR-MAD-2022-12375913], SISTE [Società Italiana di Scienze Applicate alle Piante Officinali e ai Prodotti per la Salute]; European Commission [EUROPEAID/173691/DD/ACT/XK Nanokos].

